# What is the suitability of clinical vignettes in benchmarking the performance of online symptom checkers? An audit study

**DOI:** 10.1101/2021.07.29.21261320

**Authors:** Austen El-Osta, Iman Webber, Aos Alaa, Emmanouil Bagkeris, Saba Mian, Mansour Sharabiani, Azeem Majeed

**Affiliations:** Imperial College London Self-Care Academic Research Unit (SCARU); Imperial College London, National Health & Lung Institute; Imperial College London, Department of Primary Care & Public Health

## Abstract

**Objective:** To assess the suitability of primary care vignettes in benchmarking the performance of online symptom checkers

**Design:** Observational study using publicly available, free online symptom checkers

**Participants:** Three symptom checkers (Healthily, Ada and Babylon) that provided consultations in English. 139 standardized patient vignettes were compiled by RCGP. Three independent GPs interpreted the vignettes to arrive at a “Gold Standard” consisting of 3 dispositions and divided into one of three categories of triage urgency: (1) emergency care required, (2) primary care required and (3) self-care.

**Main outcome measures:** Six professional non-medical and lay inputters simulated 2774 standardized patient evaluations using 3 online symptom checkers (OSC). We recorded when OSC provided a triage recommendation and whether it correctly recommended the appropriate triage recommendation across three categories of triage urgency (emergency care, primary care or self-care). We collected data on whether the solution appeared within the first 3 dispositions in each of the standards across 2774 standardized patient evaluations.

**Results:** When benchmarked against the Gold Standard, Healthily provided an appropriate triage recommendation 61.9% of the time compared to 45.3% and 42.4% of the time for Babylon and Ada respectively. There was poor agreement between OSC consultation outcome and Gold Standard dispositions. When compared to the Gold Standard, Healthily gave an unsafe “under-triage” recommendation 28.6% of the time overall across the three categories compared to 43.3% for Ada and 47.5% for Babylon (P<0.001).

**Conclusions:** OSCs recommended ‘very unsafe’ triages only <4% of the time suggesting that the online consultation tools are generally working at a safe level of risk. Primary care vignettes are a helpful tool to support development of OSC, but not ideally suited to benchmark the performance of different OSC. Real-world evidence studies involving general practice are recommended to benchmark the performance of OSC in the community setting.

**Strengths and limitations of this study:**

- 139 independently created primary care vignettes covering 18 subcategories of primary care were used to benchmark the performance of three online symptom checkers using 2774 unique patient simulations
- A gold standard for each primary care vignette was derived using GP roundtables and single blinded testing
- We investigated the extent that different inputters using the same vignette and online symptom checker received differing consultation outcomes and triage recommendations
- We developed an accuracy matrix to objectively monitor online symptom checker consultation outcome and the safety of the triage recommendation
- Limitations included a different number of inputters to simulate patients across the three online symptom checkers tested

## INTRODUCTION

In the USA, over a third of adults self-diagnose their conditions using the internet, including queries about urgent (i.e., chest pain) and non-urgent (i.e., headache) symptoms(1, 2). The main issue with self-diagnosing using websites such as Google and Yahoo is that the users may get confused or receive inaccurate information, and in the case of urgent symptoms, the users may not appreciate the need to seek emergency care (3). In recent years, various online symptom checkers (OSC) based on algorithms or artificial intelligence (AI) have emerged to fill this gap.

OSCs are calculators that ask users to input details about their symptoms of sickness, along with personal information such as gender and age. Using algorithms or AI, the symptom checkers propose a range of conditions that fit the symptoms the user experiences. Developers promote these digital tools as a way of saving time for patients, reducing anxiety and giving patients the opportunity to take control of their own health(4-6). The diagnostic function of OSC is aimed at educating users on the range of possible conditions that may fit their symptoms. Further to present a condition outcome and give the users a triage recommendation that prioritises their health needs. The triage function of OSC guides users on whether they should self-care for the condition they are describing or whether they should seek professional healthcare support (3). This added functionality could vastly enhance the usefulness of OSC by alerting people about when they need to seek emergency support or seek non-emergency care for common or self-limiting conditions (7).

Babylon has claimed that their OSC performed better than the average doctor on a subsection of the Royal College of General Practitioners (RCGP) exam (8). This claim has been supported by an internal evaluation study (9), but the findings were later considered uncertain due to methodological concerns (10, 11). Misdiagnosis of patients with life-threatening conditions could worsen their health, especially if they are not told to seek care when they should, resulting in an increased risk of preventable morbidity and mortality. In spite of this, there has been little evidence in previous literature to suggest if OSC are harmful to patients (12, 13). However, OSC that have high false-negative rates may run similar risks if used by patients with high-risk disease such as cardiac ischaemia, pulmonary embolism or meningitis (5). With this in mind, it is extremely important that there are guidelines on robust evaluation of OSC regarding patient safety, efficacy, effectiveness and cost (5).

Very little research has been done on the performance of symptom checkers for actual patients (14-19). Equally, there is a limited number of studies that attempted to benchmark the performance of different OSC using clinical vignettes (20-26). A recent study compared the breadth of condition coverage, accuracy of suggested conditions and appropriateness of urgency advice of eight popular OSC (24), and showed that the best performing OSCs have a high level of urgency advice accuracy which is close to that of GPs and are close to GP performance in providing the correct condition in their top-3 condition suggestions OSC (24). However, it remains uncertain if clinical vignettes are ideal to investigate the accuracy and safety of OSC generally. To address this gap in knowledge, we worked in collaboration with RCGP to develop a methodology to determine if clinical vignettes were a suitable tool that can be used to benchmark the performance of different OSC.

Our approach included the creation of an independent source of vignettes from RCGP to arrive at a “Gold Standard” of medical opinion that worked from the condition to the outcome and the vignette to the outcome - as non-currently exists. The Gold Standard was used to explore the issues we faced with variable interpretations of vignettes.

### Study aim

The primary aim of this study was to assess the suitability of primary care vignettes in benchmarking the performance of online symptom checkers. The secondary aim was to benchmark the safety and efficacy of a three popular online symptom checkers (Healthily, Ada and Babylon) against current Gold Standard. Safety was defined as giving the appropriate triage recommendation (the primary outcome) relative to the current Gold Standard, whereas accuracy was defined as providing the correct outcome consultation (the secondary outcome) for each vignette. We also sought to investigate the extent that interpretations of the same vignette by different inputters could lead to different outputs using the same OSC.

## METHODS

139 primary care clinical vignettes representing 18 sub-categories of primary care **(Table 1)** were devised as illustrated in **Figure 1**. This involved a series of roundtables discussions at the Royal College of General Practitioners (RCGP) and consolidation of additional deliberations by independent GP partners to arrive at the Gold Standard. **Figure 2** shows the testing process of the online symptom checkers (OSC) by three lay and four professional non-doctor inputters using the Gold Standard vignettes. This allowed us to benchmark the performance of online symptom checkers and to determine if clinical vignettes are a suitable methodology to compare the performance of OSC.

**Table 1:**
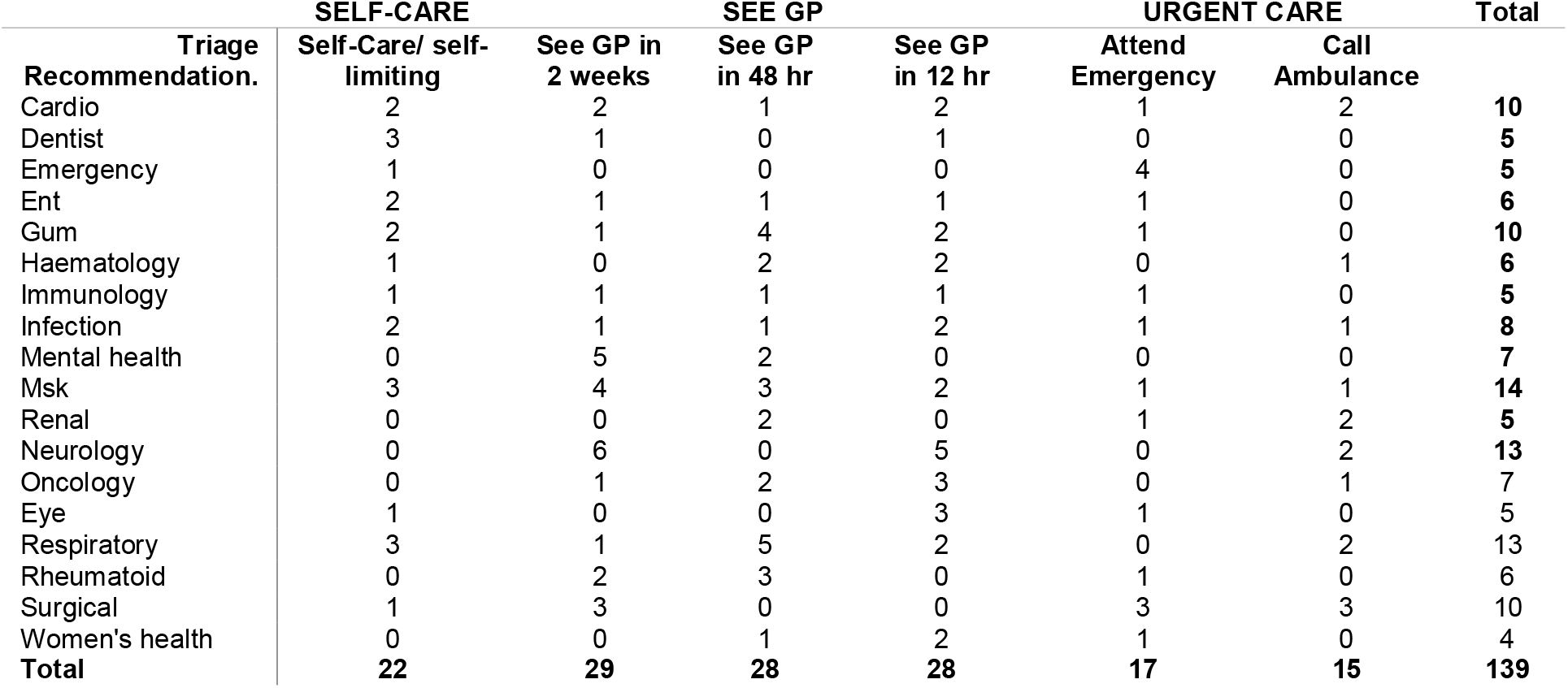
Triage recommendations for 139 vignettes across 18 subcategories of primary care.

**Figure 1:**
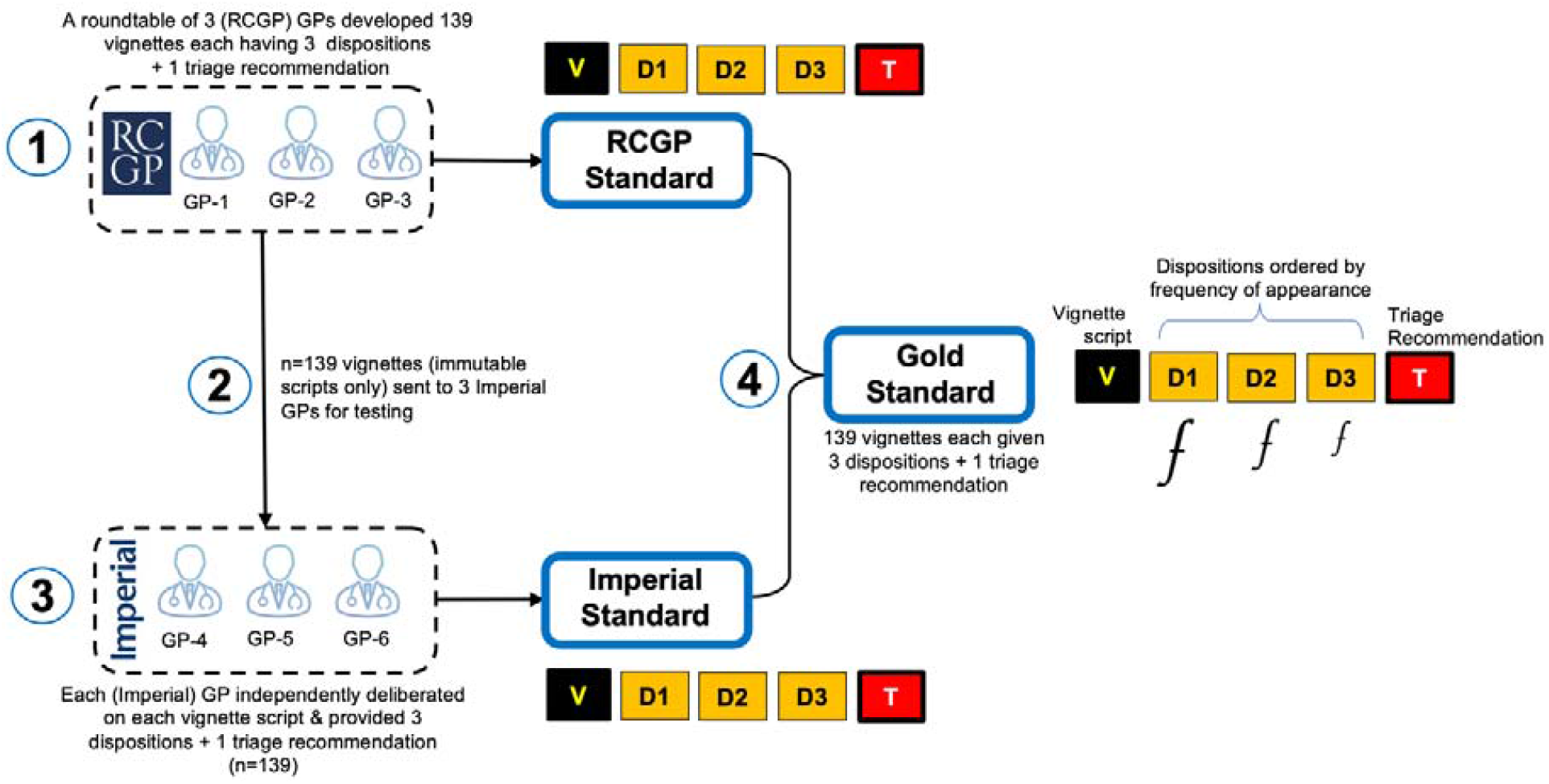
Primary care vignette creation process.

**Figure 2:**
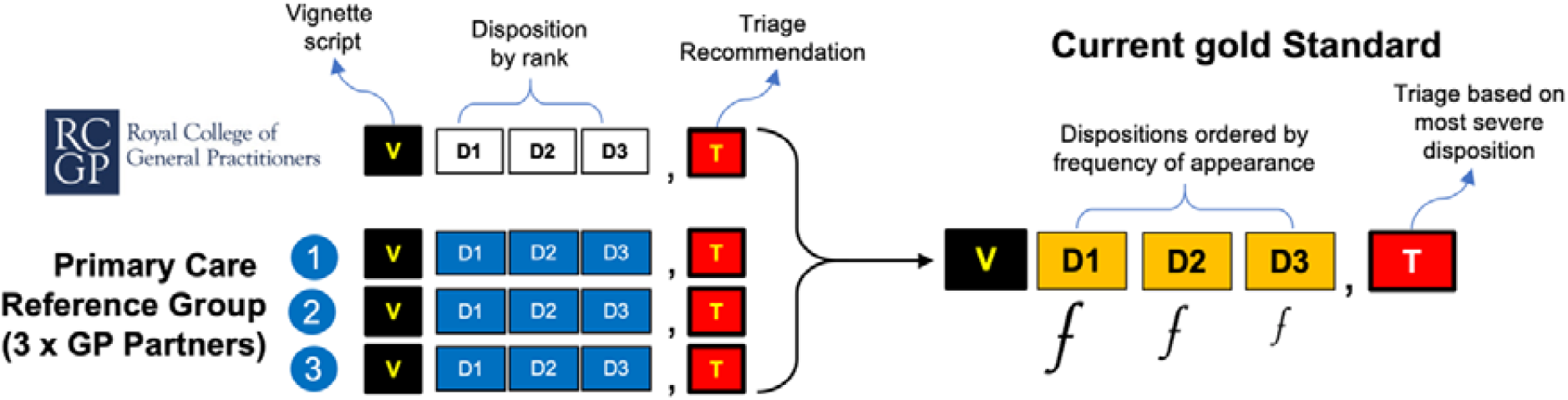
Creation of the Gold Standard using RCGP and independent GP roundtables.

### Clinical vignette creation

A roundtable of experienced General Practitioners (GP) affiliated to the RCGP supported the development of 139 primary care vignettes. The vignettes were designed to include both common and less-common conditions relevant to general practice, including clinical presentations and conditions representing 18 sub-categories of primary care **(Table 1)**.

Most of the clinical vignettes described new presentations by adults (18yr-65yr) but assumed that none of the patients were pregnant or had any prior or existing long-term conditions such as diabetes, hypertension, cardiovascular disease, terminal illness or other co-morbidity. Each vignette was created with a list of 3 reasonable condition outcomes, and an appropriate triage recommendation arrived at through the majority decision of the vignette creation RCGP Roundtable.

### Vignette characteristics

Each vignette (V) script was assigned with three dispositions (D1-D3) describing the most likely ‘diagnosis’ in D1, and the least likely in D3 **(figure 1)**. Each vignette was also given a triage recommendation (T) which was based on the most likely outcome (D1), and could assigned to any one of three categories: (1) Self-Care (i.e., see pharmacist, self-limiting condition or self-care); (2) Primary Care (i.e., see GP/Doctor in 12hr, 48hr or 2 weeks), or (3) Emergency Care (i.e., seek emergency treatment, or call ambulance). The characteristics of each vignette can be summarised in a simple 5-item cellular configuration illustrating the arrangement of D1, D2 & D3 and the T for each V **(figures1 & 2)**.

### External review of vignette by independent GPs

We provided the vignette scripts to 3 GPs affiliated to Imperial College London that had no connection with any of the OSC providers. The list of 139 vignettes were provided without the diagnosis or triage recommendations proposed by the RCGP roundtable. We asked the GPs to independently deliberate on each vignette and record up to three dispositions and one triage recommendation. The triage recommendation was again based on the most serious disposition for each vignette. This resulted in the genesis of an alternative standard to the one provided by the RCGP **(figure 1)**. This so called ‘External GP (Imperial) Standard’ was then considered in light of the RCGP Standard and both were consolidated to arrive at the Current Gold Standard **(figure 2)**. The Gold Standard was used to benchmark the performance of all three online symptom checkers.

### Synthesising the Gold Standard

The vignettes developed by the RCGP included a set of 3 dispositions and a triage recommendation as deemed suitable by the RCGP roundtable. Working on the assumption that the vignette script provided by the RCGP was immutable, the Gold Standard dispositions and triage recommendation for each vignette was synthesised by consolidating the deliberations of three additional GP Partners with the original RCGP Standard. The Gold Standard dispositions (D1, D2 & D3) for each vignette are therefore a synthesis from up to 12 dispositions; 9 dispositions in total from the 3 external GPs, and 3 dispositions form the original RCGP Standard **(2)**. When producing the Gold Standard for each vignette, the dispositions were ranked by frequency of appearance in the series; the dispositions that were most frequent appeared in D1, and subsequently in D2 & D3 **(figure 2)**. There were occasions when only D1 & D2 had a frequency of 2 or above. On occasion that D2 & D3 had frequencies of just 1 each, it was not possible to order them objectively without introducing bias. In these instances, we reverted to the original RCGP Standard to assign D2 & D3. The triage recommendation for each vignette in the Gold Standard was set to the most severe disposition in the series.

### Benchmarking OSC performance using 139 primary care vignettes

The 139 vignettes were used by both lay and professional non-doctor inputters to benchmark the performance of three online symptom checkers (Healthily, Ada and Babylon). Benchmarking was done for each OSC against all three standards: (1) the original RCGP Standard, (2) the External GP (Imperial) Standard, and (3) the Gold Standard (**figure 3**).

**Figure 3:**
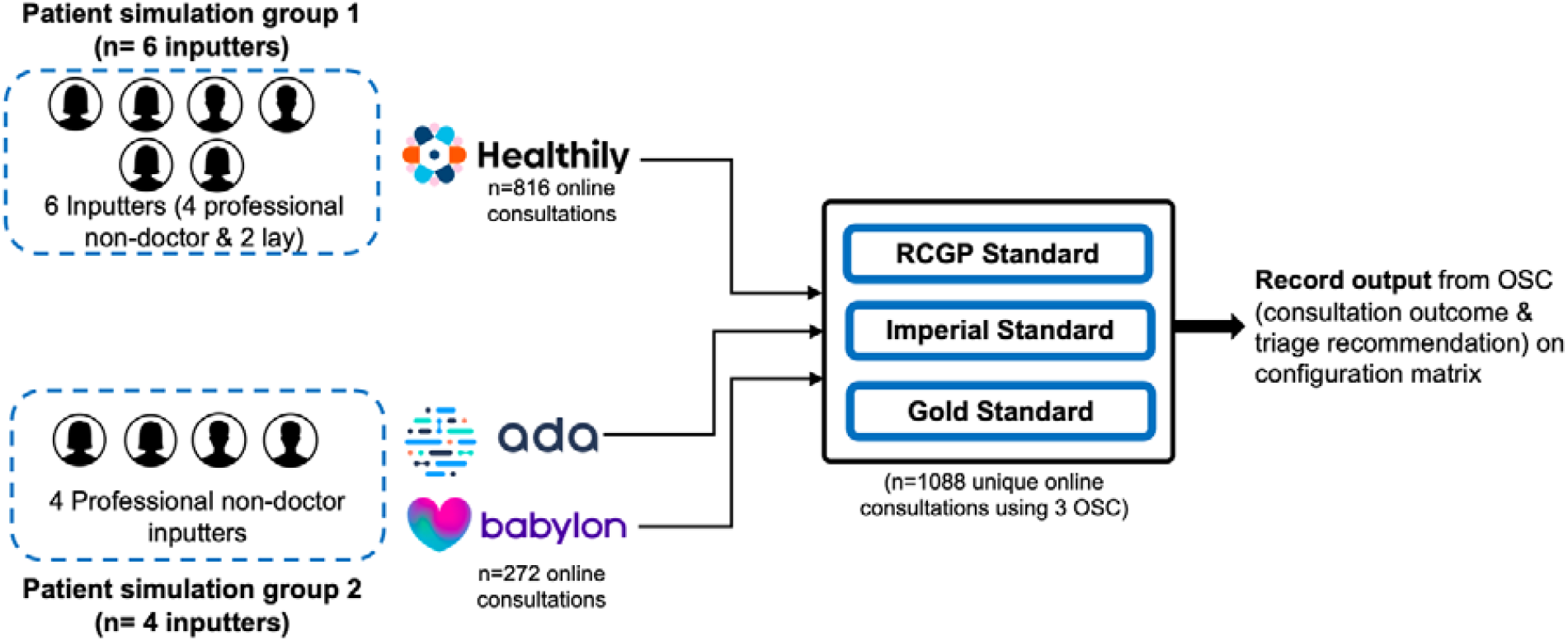
Using lay and professional non-doctor inputters to benchmark the performance of three OSC against RCGP, Imperial & Gold standards.

### Patient simulation by lay and professional non-doctor inputters

Lay and professional non-doctor inputters (n=6) used the vignettes to independently record the consultation outcome and triage recommendation from Healthily, Ada and Babylon OSC. Only one professional non-doctor inputter recorded the consultation outcome and triage recommendation using Ada and Babylon OSC for all 139 vignettes. Interpretation of the vignette script was left to the individual inputters who did not have any additional information. The inputters were instructed to make the following blanket assumptions when answering the questions posed by the OSCs for each vignette: the simulated patient is a non-smoker, not pregnant, not obese, not taking medication, not diabetic, not hypertensive, has no history of heart disease asthma, cancer, cystic fibrosis or other concerning or significant medical history, with no recent (3 months) sexual activity. Inputters were instructed to not include more than three consecutive symptoms in a single answer to any questions posed by the OSCs.

### Recording output from online symptom checkers

Different OSCs may give up to 7 consultation outcomes (range= 0-3 for Healthily, 0-5 for Ada, and 0-7 for Babylon) and a triage recommendation may or may not be provided. Inputters independently simulated the patient described in each vignette and recorded the consultation outcome using an electronic OSC consultation record form. Data were collected on up to 3 keywords used during input, details of any inputted keyword recognised by OSC, the first 3 consultation outcomes (if any) provided by OSC, the triage recommendation (if any) and whether the inputter was signposted to relevant information at end.

### Consultation outcome data coding

To support with data management and analysis of output from OSC, we developed a simple framework. We assessed output parameters for each vignette using all three OSCs against the dispositions of the three standards (RCGP, Imperial and Gold standards). We assigned a three-digit numeric score to objectively characterise the level of agreement between the three dispositions (D1, D2, D3) in each standard and the consultation outcome for each vignette:

- *3: Full agreement*; Correct consultation outcome in the exact same position as the disposition cell in the RCGP standard
- *2: Partial agreement*; Correct consultation outcome, but 1 cell apart from the RCGP disposition (e.g., D1 placement is found in D2, or D2 placement found in either D1 or D3 juxtaposing cell)
- *1: Partial agreement*; Correct consultation outcome, but 2 cells apart from the RCGP disposition (e.g., D1 placement is found in D3 or vice versa)
- *0: No agreement;* Incorrect consultation outcome in any cell, and not relating to any RCGP disposition
- *9: Null, or no output provided*

The resulting 3-digit score described an output pattern that could be objectively scored and weighted to benchmark the performance of different OSCs against each vignette standard. Of the 125 possible permutation of the 3-digit score, only 59 combinations were considered logical in representing the consultation outcome by the OSC **(supplementary table 1)**. The same terminology was used to describe the level of agreement between the dispositions in each Standard and OSC main triage recommendation.

### Statistical analysis

The consultation outcomes and triage recommendations from Healthily, Ada and Babylon OSC were compared to the RCGP, Imperial and the Gold standards for each vignette **(figure 3)**. We investigated the extent that different interpretations of the same vignette by different inputters resulted in different consultation outcomes when using the same OSC (Healthily). Descriptive analysis was used to assess the accuracy and safety of Healthily (using 6 inputters), Ada (using 1 inputter) and Babylon (using 1 inputter) against all three standards (the original RCGP standard, the Imperial standard from three external GP partners, and the consolidated Gold standard). Data were expressed in frequencies, proportions and 95% Confidence intervals (CI). Pearson’s Chi-square test and Fisher’s exact test were used to determine whether there was a difference in signposting, the provision of a consultation outcome or a triage recommendation by different inputters using the same OSC and vignette. Significance was noted when p-value was <0.05. The statistical analysis was performed using StataCorp. 2019. Stata Statistical Software, Release 16.

### Patient and Public involvement

Patient and public involvement (PPI) was embedded in this project. Two lay non-doctor inputters were involved in the collection of output data from Healthily OSC.

## RESULTS

### Comparing interpretation of the same vignette by different inputters

We compared the consultation outcome from Healthily OSC of two lay inputters against the output recorded by four professional non-doctor inputters to determine the extent that individuals could interpret the same vignette differently **(table 2; figure 4)**. A significant difference was observed for consultation outcomes in disposition cells one (p<0.001) and three (p=0.03) between both type of non-doctor inputters), but not for disposition cell two (p=0.30) or the single triage option (p=0.93).

**Table2:** Consolidated Healthily output from 2 lay and 4 professional non-doctor inputters.

| Output from 2 lay inputters | Outcome 1 |  | Outcome 2 |  | Outcome 3 |  | Triage Option |  |
| --- | --- | --- | --- | --- | --- | --- | --- | --- |
|  | N | (%) | N | (%) | N | (%) | N | (%) |
| 3 (Agree) | 133 | 47.8 | 39 | 14.0 | 19 | 6.8 | 93 | 33.5 |
| 2 (Partial) | 20 | 7.2 | 0 | 0.0 | 0 | 0.0 | - | - |
| 1 (Partial) | 10 | 3.6 | 8 | 2.9 | 1 | 0.4 | - | - |
| 0 (False) | 47 | 16.9 | 5 | 1.8 | 0 | 0.0 | 120 | 43.2 |
| 9 (Null) | 68 | 24.5 | 226 | 81.3 | 258 | 92.8 | 65 | 23.4 |
| Correct (any) | 77 | 27.7 | 13 | 4.7 | 1 | 0.4 | 120 | 43.2 |
| Non-correct | 201 | 72.3 | 265 | 95.3 | 277 | 99.6 | 158 | 56.8 |
| **Output from 4 professional inputters** |  |  |  |  |  |  |  |  |
| 3 (Agree) | 207 | 37.2 | 64 | 11.5 | 19 | 3.4 | 188 | 33.8 |
| 2 (Partial) | 31 | 5.6 | 0 | 0.0 | 3 | 0.5 | - | - |
| 1 (Partial) | 32 | 5.8 | 26 | 4.7 | 4 | 0.7 | - | - |
| 0 (False) | 145 | 26.1 | 17 | 3.1 | 2 | 0.4 | 241 | 43.3 |
| 9 (Null) | 141 | 25.4 | 449 | 80.8 | 528 | 95.0 | 127 | 22.8 |
| Correct (any) | 208 | 37.4 | 43 | 7.7 | 9 | 1.6 | 241 | 43.3 |
| Non-correct | 348 | 62.6 | 513 | 92.3 | 547 | 98.4 | 315 | 56.7 |

**Figure 4:**
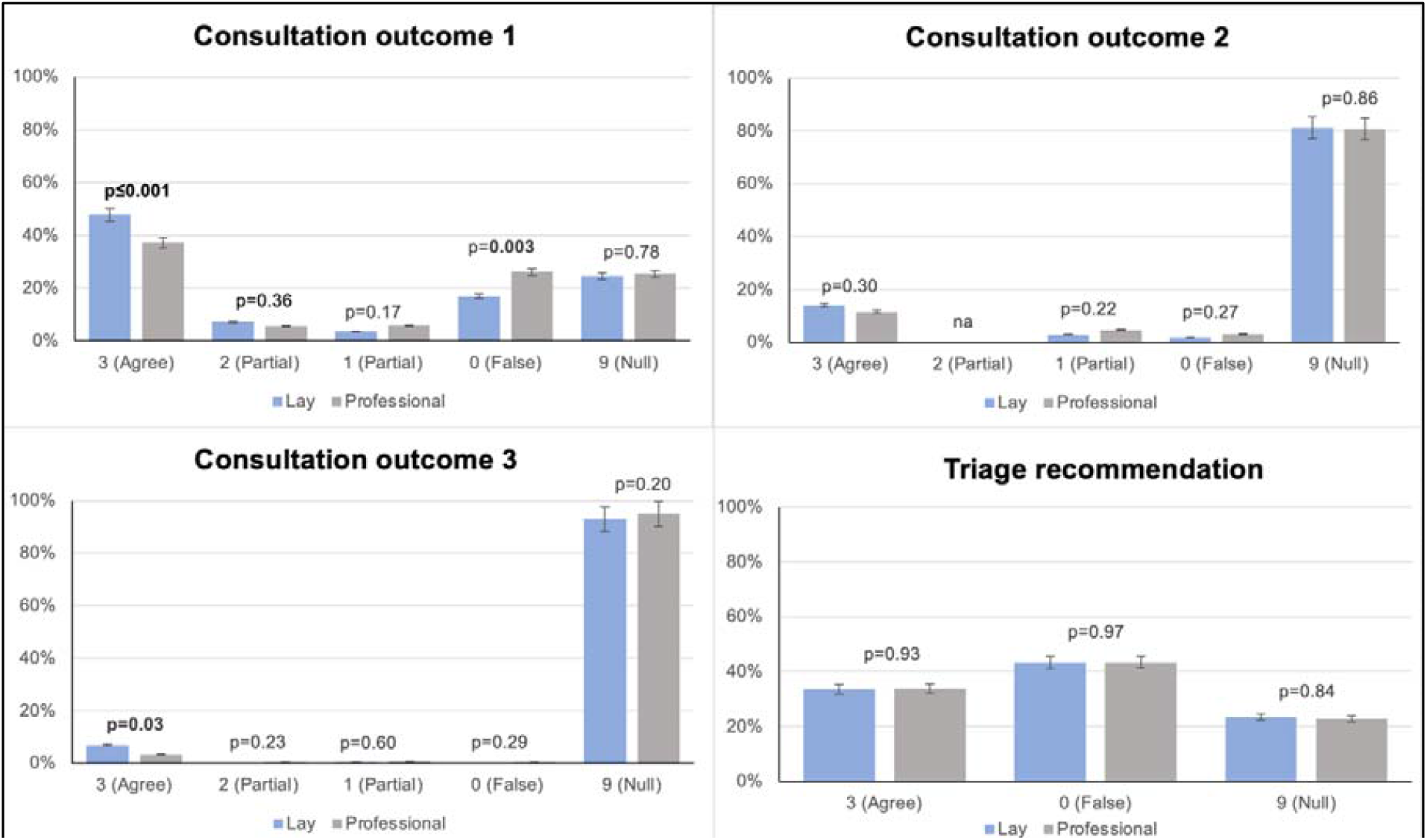
Levels of agreement between different inputters.

### Signposting at end of consultation outcome

There was no significant difference in signposting between professional inputters when using Healthily OSC (p=0.23). However, there was a significant difference between the two lay inputters (p<0.001), and between the professional (n=4) and lay (n=2) inputters when compared as a group (p<0.001). There was significant variation between inputters with respect to whether the same OSC (Healthily) provided signposting at the end for the same vignette, regardless of whether the simulation resulted in a triage recommendation or not but when there was no triage recommendation (p<0.001; **table 3**). The difference disappeared when no triage recommendation was provided (p=0.21).

**Table 3:** Level of agreement between inputters when using the same OSC to simulate the patient described in each vignette script.

| Inputter | Signposting pathway | Regardless of whether they got a triage recommendation or not |  | When there was no triage recommendation |  |
| --- | --- | --- | --- | --- | --- |
|  | Yes/No | N | (%) | N | (%) |
| Professional inputter 1 | No | 12 | (8.6) | 0 | (0.0) |
|  | Yes | 127 | (91.4) | 11 | (100.0) |
| Professional inputter 2 | No | 18 | (12.9) | 0 | (0.0) |
|  | Yes | 121 | (87.1) | 0 | (0.0) |
| Professional inputter 3 | No | 9 | (6.5) | 2 | (14.3) |
|  | Yes | 130 | (93.5) | 12 | (85.7) |
| Professional inputter 4 | No | 17 | (12.2) | 1 | (33.3) |
|  | Yes | 122 | (87.8) | 2 | (66.7) |
| Lay inputter 1 | No | 8 | (5.7) | 0 | (0.0) |
|  | Yes | 131 | (94.2) | 0 | (0.0) |
| Lay inputter 2 | No | 43 | (30.9) | 0 | (0.0) |
|  | Yes | 96 | (69.1) | 0 | (0.0) |
| Total | No | 107 | (12.8) | 3 | (10.7) |
|  | Yes | 727 | (87.2) | 25 | (89.3) |

### Assessing the level of agreement between external GP partners when assigning dispositions and triage recommendations for each vignette script

When GPs deliberated on the vignette scripts provided to arrive at the Imperial Standard, we found low (36.4%) agreement for triage recommendations for ‘self-care’ between the RCGP and Imperial Standard, moderate (58.8%) agreement for ‘see doctor’ triage recommendations, and higher (68.8%) agreement for vignettes suggesting emergent care **(table 4)**.

**Table 4:** Level of agreement for triage recommendation between RCGP and Imperial standards.

|  | Total *n* of Vignettes in triage category (original set) | Triage agreement between Imperial & RCGP | % agreement |
| --- | --- | --- | --- |
| Self-Care | 22 | 8 | (36.4%) |
| Primary Care | 85 | 50 | (58.8%) |
| Emergency | 32 | 22 | (68.8%) |
| TOTAL | 139 | 80 | (57.6%) |

**Table 5** illustrates the level of agreement between Imperial GPs against the RCGP Standard for disposition cell 1 (D1). There was unanimous agreement between the deliberations of the three external GPs for the preferential diagnosis in disposition one (D1) and D1 of RCGP Standard only 32.4% of the time, and partial agreement for D1 25.9% and 14.4% (40.3% total) on D1 where only two or one GPs agreed with RCGP in that same order (**table 5**). There were only 38 instances (27.3%) where none of the three external GPs proposed a diagnosis that matched D1 of the RCGP Standard.

**Table 5:**
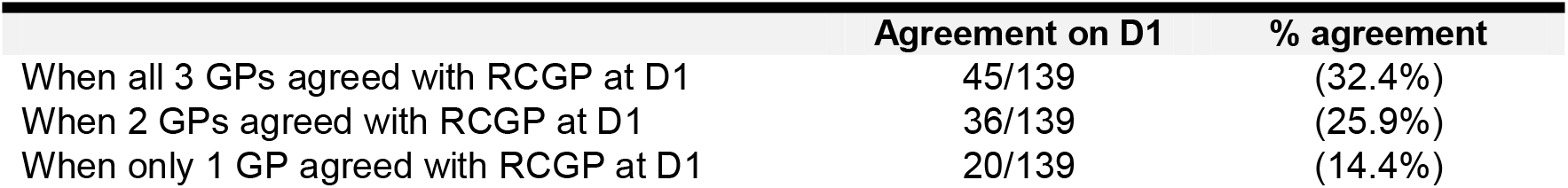
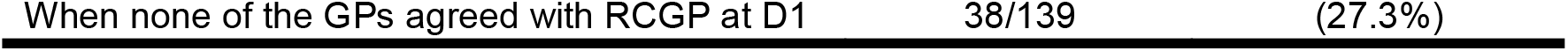
Level of agreement between three external GP partners regarding disposition 1 (D1)

There was a good level of agreement overall (74.6%) between the consolidated triage recommendations of the three external GPs (the Imperial Standard) and the RCGP Standard **(table 6)**. There was also a good level of agreement (72.2%) between the consolidated Imperial Standard when assessed against the RCGP Standard at disposition cell one (D1), 31.2% at D2 and 12.5% at D3 **(table 6)**.

**Table 6:**
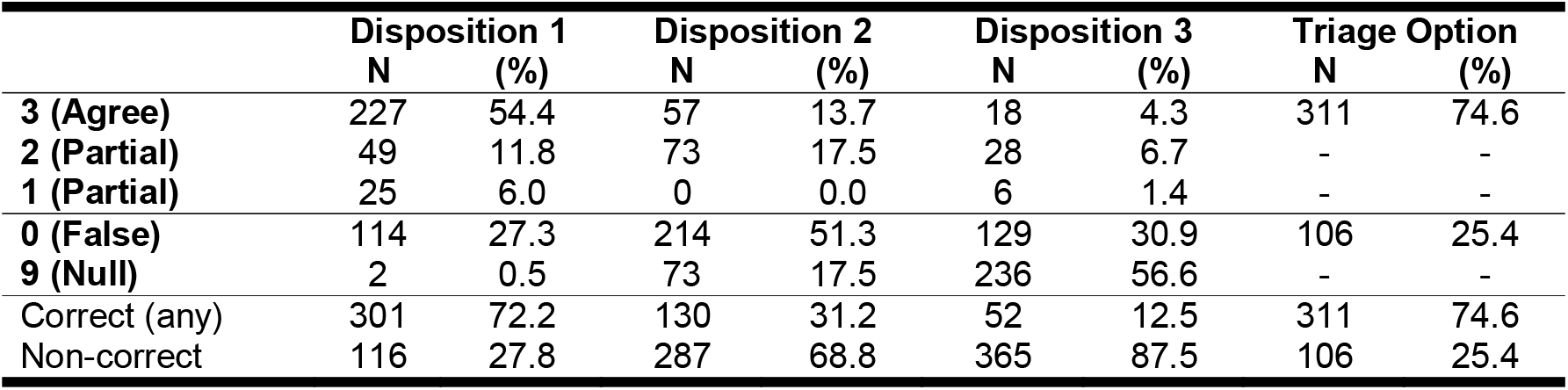
Levels of agreement between consolidated Imperial Standard & RCGP Standard.

### Benchmarking performance of three OSC against current Gold Standard

We used one professional non-medical inputter to engage with Ada and Babylon OSC, and wanted to test the methodological approach with different symptom checkers to see if the same issues arose and to benchmark performance. Ada consistently performed better than Healthily and Babylon in providing the correct consultation outcome in D1, D2 and D3. The correct consultation outcome for Ada against the RCGP Standard at any disposition was 54.0% compared to 37.4% for Healthily and 28.1% for Babylon (**supplementary table 2;** p<0.001). Contrary to Ada, the performance of Healthily and Babylon OSC improved significantly when comparing the level of agreement for D1 to the RCGP and the Gold standards (**supplementary table 2**; p<0.001). Ada nearly always provided at least one consultation outcome at D1 (range (97.1%, 100%) for all patient simulations, whereas Babylon and Healthily only provided a consultation outcome some of the time (Healthily range (74.6%, 75.5%); Babylon range (71.9%, 77.7%)) for D1 respectively; s**upplementary table 2**).

### Accuracy of triage recommendations

In benchmarking against the original RCGP standard, Healthily provided an appropriate triage recommendation 43.3% (95% CI 39.2%, 47.6%) of the time, whereas Ada and Babylon were correct 61.2% (95% CI 52.5%, 69.3%) and 57.6%, (95% CI 48.9%, 65.9%) of the time respectively (p<0.001). However, the situation was reversed when OSCs were benchmarked against the Gold Standard as Healthily provided a congruent triage recommendation 61.9% (95% CI 57.7%, 65.9%) of the time, compared to 42.4% (95% CI 34.1%-54.1%) and 45.3% (95% CI 36.9%, 54.0%) of the time for Ada and Babylon respectively (p<0.001; **figure 5**).

**Figure 5:**
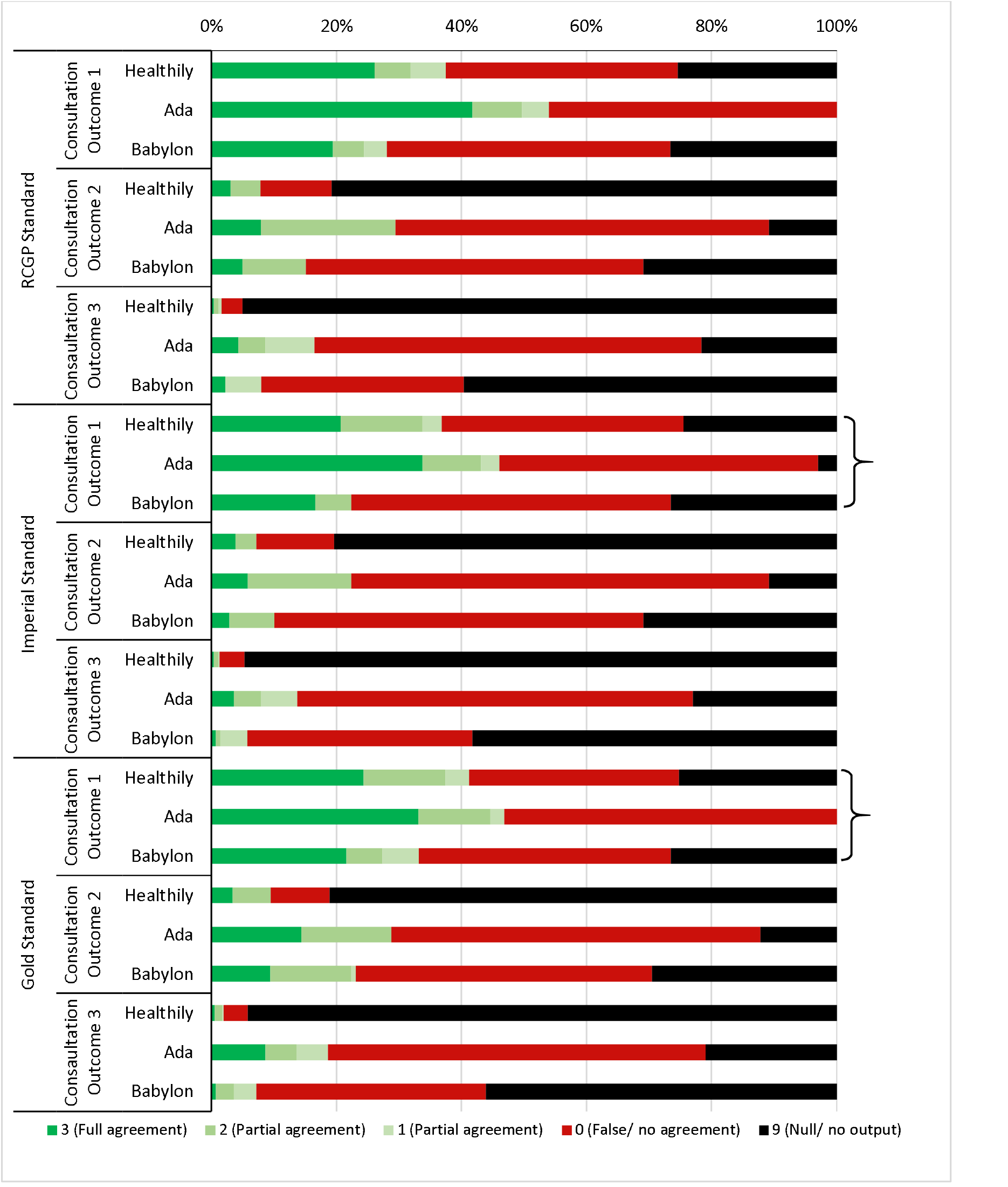
Comparing the accuracy of consultation outcomes at D1, D2 and D3 between three OSC using 139 vignettes against three standards.

### Triage safety

When compared against the Gold Standard, Healthily made triage recommendations that were deemed unsafe 0%, 4.3% and 24.3% of the time for vignettes describing a symptom that required self-care, primary care and emergent care respectively **(supplementary table 3)**. This percentage of unsafe triages contrasted to 0%, 18.4% and 24.9% for Ada and 0%, 9.2% and 38.2% for Babylon in that same order. When compared to the Gold Standard, Healthily gave unsafe triage recommendations only 28.6% of the time overall across the three categories compared to 43.3% for Ada and 47.5% for Babylon (P<0.001; **supplementary table 3**). Healthily was significantly more likely to recommend safe triages for vignettes indicating primary care than Ada (p<0.001), but the difference did not reach the level of significance when compared to Babylon (p=0.07). Similarly, Healthily was significantly more likely to recommend safe triages for vignettes indicating emergent care than Ada (p<0.001), but the difference did not reach the level of significance when compared to Babylon (p=0.10), f**igure 6**.

**Figure 6:**
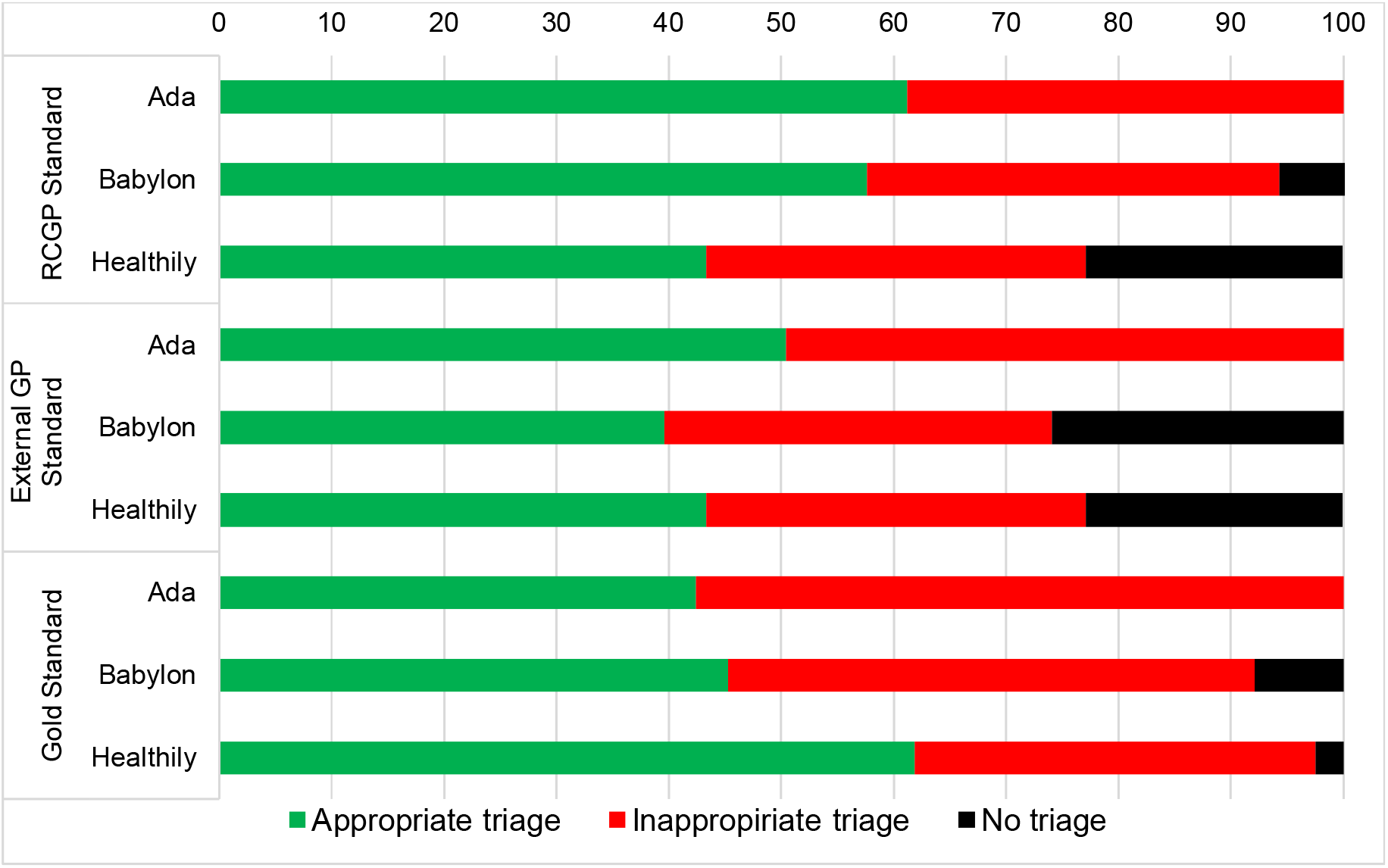
Accuracy of triage recommendations from OSCs against three standards.

**Figure7:**
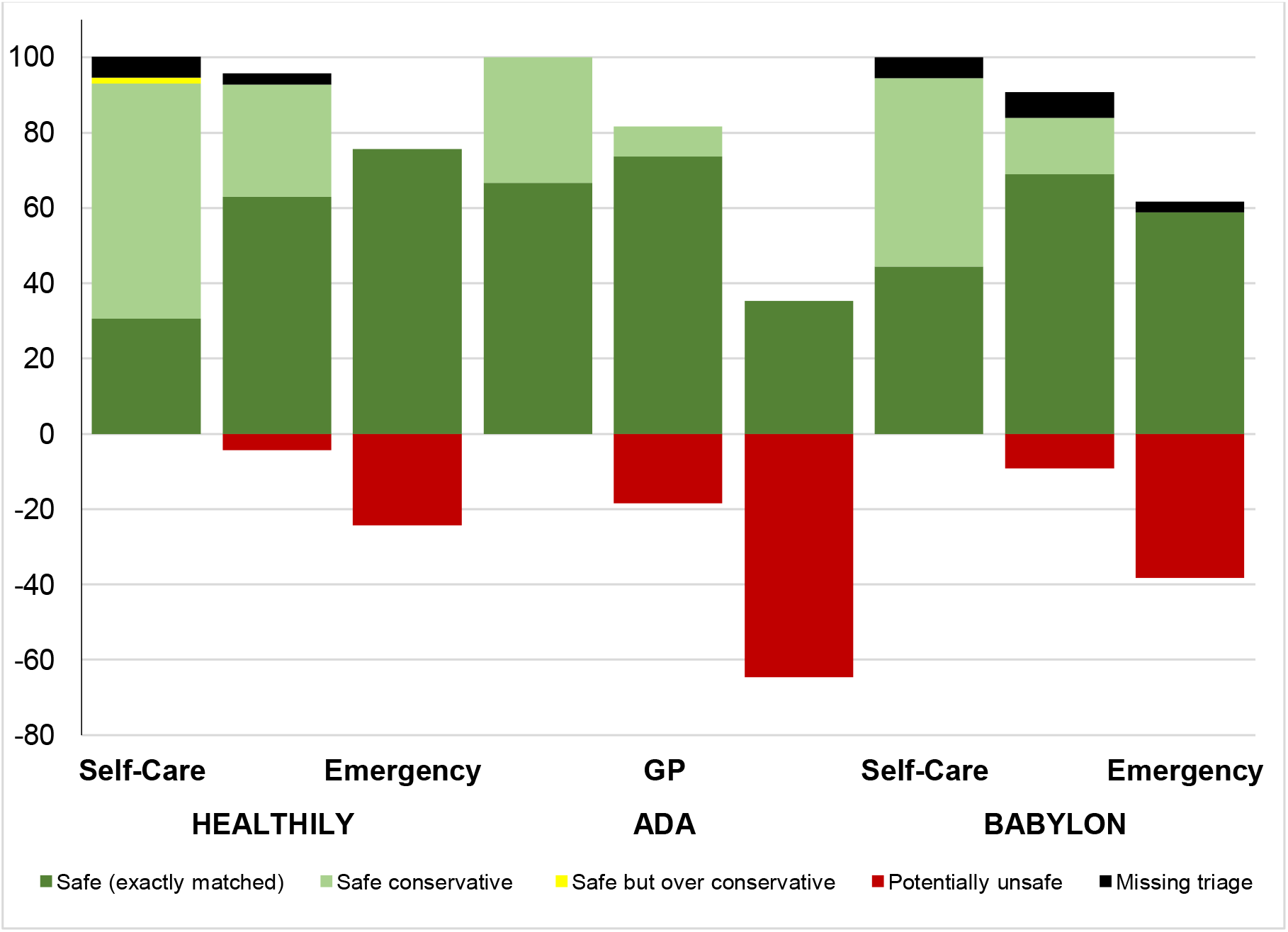
Accuracy of urgency of triage recommendations displayed as a stacked bar chart against on the gold standard triage. For Healthily, four professional non-doctor inputters recorded the data, whereas only one inputter recorded data for Ada and Babylon online symptom checkers

## DISCUSSION

This is the first study which directly sought to assess the suitability of clinical vignettes in benchmarking the performance (accuracy and safety) of OSC. To facilitate this, an independent series of primary care vignettes provided by the RCGP describing patient scenarios and symptoms across 18 sub-categories of primary care was needed. This new set was used to compare baseline performance of OSC against two iteratively refined and consolidated standards arising from further deliberations by an additional roundtable of independent general practitioners.

Our study showed significant variability of medical opinion depending on which group of GPs considered the vignette script, whereas consolidating the output of two different GP roundtables (RCGP and Imperial) resulted in a more refined third iteration representing the Gold Standard which by definition most accurately describes the most appropriate ‘diagnoses’ conferred by the vignette script. The different qualities of each standard suggests that clinical vignettes are not an ideal tool for benchmarking the accuracy of OSC since performance will always be related to the nature and order of the dispositions which we have shown can differ significantly between each standard depending on the approach and levels of input form GP roundtables. By extension, the Gold Standard can always be improved by consolidating the deliberations of a wider range medical opinion until saturation is reached and a final consensus emerges.

Another key factor that may impact the benchmarking of OSC using clinical vignettes even on occasion that a refined Gold Standard is used is related to the inputter’s inability to answer truthfully to all questions that could be asked during the online consultation process. This inherent methodological limitation necessitates the use of blanket assumptions (e.g., not pregnant, did not have any recent sexual activity etc.,) which could lead to a different consultation outcome to what would be presented if a real patient who was experiencing the symptoms was the inputter. For example, the significant difference in OSC output from different inputters suggests that the wording of some items needs to be revised to reduce the likelihood of divergent interpretations of the same vignette script. It is inevitable that different people will use different words to describe their conditions, necessitating the use of machine learning to render OSC capable of understanding multiple different descriptions of the same problem. Further, the vignette script is necessarily limited in the number of words, and even if the description and context were expanded it may still not capture all the information necessary to simulate how a real patient may engage with the same OSC. This inherent limitation was illustrated by the finding that different inputters arrived at different consultation outcomes even when using the same vignette script and the same OSC.

The primary outcome measure in our study was the triage recommendation of the vignette. From the perspective of the RCGP Roundtable, the triage recommendation was always based on the first (most likely) disposition (i.e., D1) for each vignette. This was contrasted in the Gold Standard which assigned a triage recommendation based on the most severe disposition for each vignette regardless of the most likely outcome. Recommending a triage option that is based on the ‘worst case scenario’ (above a certain likelihood threshold) is usually recommended as it ensures patient safety. For example, if dispositions 1, 2 & 3 for a vignette were Indigestion, Costochondritis and Heart attack, the triage will be for ambulance based on D3. This is clearly the safer of the two options and the one adopted by Healthily, whereas Ada and Babylon provide a triage consultation based on the first disposition in the series. This may explain why the performance of Healthily, and to a lesser extent Babylon, in recommending safe triages improved when compared to Ada which routinely makes the triage recommendation based on the most likely diagnosis (i.e., D1) as opposed to the most serious disposition in the series.

Pertinently, the independent deliberations of external GPs only agreed with RCGP dispositions on average 72% of time and on triage 74% of the time, but this did not mean that any of the GPs were wrong. When GPs diagnose, they assess probable risk and then investigate implying that primary care assessment is not binary; there is often not a correct answer but rather a series of options that could be explored with the patient to help resolve the symptoms and treat the condition using evidence-based decision refined over time including the use of further tests. By contrast, the provenance of a clinical vignette starts from the condition and builds a story, whereas conversely GPs and OSCs start with the story and then work towards a probable condition. Often, as we saw following the independent deliberations of three independent GPs, there are many possible conditions in the “area” a vignette might point towards. We found that often the OSC and the independent GPs were in the right area but not precisely “correct”. This demonstrates why any claims that an OSC can “diagnose” need to be challenged, since GPs do not diagnose and therefore OSCs cannot. Diagnosis can only come after testing and verification of the initial hypothesis, and accurate diagnosis usually includes other aspects such as imaging, pathology results involving the use of point-of-care and other near patient testing procedures.

Our study showed that Healthily performance improved by 6% in accuracy and by 18.2% in making an appropriate triage recommendation when benchmarked against the Gold Standard in comparison to the original RCGP Standard. On the whole, OSC recommended ‘very unsafe’ triages <4% of the time and this suggests that the online consultation tool is generally working at a safe level of probable risk. The OSCs benchmarked appeared to be ‘safe’ overall as they more frequently made an appropriate triage recommendation or signposted the user to the more urgent triage category as opposed to the other way around. It is reasonable to expect OCs to be ‘risk averse’ since these decision support tools arrive to a conclusion with limited data and without human interaction (27).

Our audit study had a number of limitations, including that the original RCGP dispositions and appropriate triage recommendations for each vignette offered a baseline for assessment but could not be considered as the Gold Standard prior to further validation and input from an external roundtable of independent GP partners. We also demonstrated a significant variation in how each of the 6 non-doctor inputters interpreted each vignette-thus often arriving at different outcome consultations (and to a lesser extent) triage recommendations when using the same online tool. We addressed this limitation by consolidating the outputs from the four professional non-medical inputters. In spite of some limitations, the framework and pragmatic methodology used to support the objective development of the Gold Standard consisting of 139 vignettes with congruent dispositions and triage recommendation were suitable to benchmark the performance of online consultation tools. We acknowledge also that the Gold Standard can be developed further by inviting input from a larger number of general practitioners. Further work is indicated to refine the wording of some vignettes since there is a large variation in how different inputters could interpret each item leading to different consultation outcomes and triage recommendation (the main output parameters) when using the same online symptom checkers.

There are a number of person-centred and policy implications for the use of OSC. For example, access to healthcare is a major issue and this has become more pronounced since the advent of the Covid-19 pandemic (28). Improving access to primary care and/or pre-primary care health advice is expected to reduce pressure on urgent and secondary care services, and this is a main driver for the use of a safe and effective OSCs. The widespread diffusion and use of OSC with added functionality can help empower individuals, improve health literacy levels through microlearning (29), and promote individual self-care capability and the rational use of products and services. This applies especially for OSC that signpost users to relevant information that could help them determine possible next steps regardless of whether or not the OSC provided a triage recommendation or not. At this stage in their development, OSC must be risk averse by avoiding under-triage where patients are directed to a less urgent service. This may have a negative impact on health service resources in that it may result in unnecessary use of urgent or emergency health providers, but may equally result in an earlier diagnosis and appropriate treatment of medical conditions which reduces morbidity, mortality and overall costs in the long term. The correct use of OSC may also decrease the high demand on primary care providers and this utility is especially welcome since the workload for GPs in the UK has increased by 62% from 1995 to 2008 (30), whereas there has been very little or no increase in the number of GPs per 1,000 population (31).

## Conclusion

Clinical vignettes are a helpful tool to benchmark the performance of OSC for research and development purposes, but inherent limitations render them largely unsuitable to compare performance between different OSCs against the Gold Standard. This study showed that online consultation tools are already working at a safe level of probable risk, but further work is recommended to cross-validate the performance of OSC against real-world test case scenarios using real patient stories and interactions with GPs as opposed to using artificial vignettes only which will always be the single most important limitation to any cross-validation study. It is also essential that these tools do not exacerbate the “digital divide” and increase health inequalities in groups such as the poor, ethnic minorities and the elderly (32).

## Data Availability

No additional data are available

## Acknowledgements

The authors thank Ms Noor Jawhar, Ms Christina Pillay and Mr John Norton for PPI input and for testing the symptom checkers, and Drs Aisha Newth, Prakash Chatlani, David Mummery and Benedict Hayhoe for clinical support.

## Author Contributors

All authors provided substantial contributions to the conception (AEO, IW, AA, EB, AM), design (AEO, EB, IW, AA), acquisition (IW, AA, ER), and interpretation (EB, IW, HM, MS) of study data and approved the final version of the paper. AEO took the lead in planning the study with support from co-authors. EB carried out the data analysis with support from AA and MS. AEO is the guarantor.

## Data sharing statement

No additional data are available

